# Prevalence and Determinants of Hypertension Among People Living with HIV on Antiretroviral Therapy: A Cross-Sectional Study at Mwananyamala Hospital, Tanzania

**DOI:** 10.1101/2025.08.12.25333448

**Authors:** Meshack Morice, Jacqueline Mgumia

## Abstract

**Background:** Hypertension is an increasingly recognized comorbidity among people living with HIV (PLHIV) receiving antiretroviral therapy (ART), contributing substantially to cardiovascular morbidity. As life expectancy among PLHIV improves, understanding both traditional and HIV-related determinants of hypertension is essential. This study assessed the prevalence, associated factors, and hypertension knowledge among PLHIV on ART in Tanzania.

**Methods:** A hospital-based cross-sectional study was conducted among 296 PLHIV on ART for more than six months at Mwananyamala Regional Referral Hospital. Data were collected using structured questionnaires and clinical records. Blood pressure was measured according to American Heart Association guidelines. Logistic regression analyses were performed to identify factors independently associated with hypertension at p<0.05.

**Results:** The prevalence of hypertension was 32.8%. Knowledge of hypertension was moderate (mean= 53.9%), with only 10.8% correctly identifying normal blood pressure levels. Age ≥55 years (aOR = 4.786, p < 0.001) and cigarette smoking (aOR = 6.159, p = 0.020) were associated with greater risk of hypertension. Long duration since HIV diagnosis (>10 years: aOR = 3.090, p = 0.011) was also associated with increased odds of hypertension, whereas shorter ART duration (<5 years: aOR = 0.106, p = 0.040) was associated with lower risk.

**Conclusions:** Hypertension is prevalent among PLHIV on ART in Tanzania, influenced by both traditional and HIV-related factors. Integrating hypertension screening and education within HIV care services is essential. Strengthening community engagement in chronic disease screening can enhance early detection, improve awareness, and promote preventive behaviours, ultimately reducing the dual burden of HIV and non-communicable diseases.

## INTRODUCTION

Hypertension, defined as blood pressure ≥140 mmHg systolic or ≥90 mmHg diastolic or use of antihypertensive medication, is a growing public health concern globally, affecting a quarter of individuals living with HIV in the previous decade and contributing to cardiovascular diseases (CVD), stroke, heart failure, eye problems (blindness) and other health-related issues[1–4] . Among people living with HIV (PLHIV), the advent of antiretroviral therapy (ART) has transformed HIV into a manageable chronic condition, significantly improving survival rates [5–7]. However, this prolonged survival has shifted morbidity patterns from opportunistic infections to non-communicable diseases, notably hypertension [8]. Studies suggest PLHIV on ART face a higher hypertension risk than the general population, potentially due to HIV-induced inflammation, ART side effects, and traditional risk factors such as alcohol drinking and cigarette smoking [9].

In 2020, global prevalence of hypertension among PLHIV on ART was reported to be 23.6% [10], with the prevalence in Sub-Saharan Africa ranging from 19.6% to 53% [11, 12]. The development of hypertension in PLHIV is multifactorial. With traditional risk factors such as age, obesity or overweight, excessive alcohol consumption, cigarette smoking, sedentary lifestyle, and family history of hypertension having been consistently associated with increased hypertension risk [1, 9, 10]. These risk factors are often more prevalent or more impactful among PLHIV due to intersecting socioeconomic, behavioural, and clinical challenges. Beyond traditional risk factors, HIV-related factors have also been associated with changes in blood pressure among people living with HIV (PLHIV). These factors include CD4 cell count, duration of HIV infection, and duration of antiretroviral therapy (ART) use. These blood pressure changes may result from prolonged immune activation and chronic inflammation caused by HIV, as well as ART-induced metabolic changes [9, 12, 13].

In Tanzania, where HIV prevalence declined from 7% in 2003 to 4.4% in 2023 among adults aged 15–49 [14, 15], hypertension among PLHIV remains understudied, yet it represents a growing public health concern even in the general population. Recent nationally representative data from the 2022 Tanzania Demographic and Health Survey indicate that the prevalence of hypertension among adults aged 15-49 years is 11%, and it is strongly associated with increasing age and overweight/obesity [16]. Another study reported a 34.6% prevalence of hypertension among PLHIV, suggesting a substantially higher burden in this population [17]. However, evidence on HIV-related factors (i.e., CD4 count, ART duration, HIV staging, and HIV diagnosis duration) and hypertension risk is inconsistent, with some studies finding no association [11, 18, 19], while others report strong links [12, 20–22]. Knowledge gaps about hypertension among PLHIV further complicate prevention efforts. Limited awareness of hypertension [22] hinders behavioural interventions, yet few studies in Tanzania have assessed this. Addressing these gaps is critical, as hypertension is a primary contributor to CVD, a leading cause of mortality globally [1]. Given the significant burden of hypertension among PLHIV, this study aimed to evaluate the determinants of hypertension among PLHIV on ART, examining risk factors and hypertension knowledge to inform targeted interventions.

## MATERIALS AND METHODS

### Study Design and Setting

This quantitative, hospital-based cross-sectional study was conducted at Mwananyamala Regional Referral Hospital (MRRH), a major referral facility in Kinondoni District, Dar es Salaam, Tanzania. MRRH’s Care and Treatment Clinic (CTC) records approximately 26,000 total patient visits per year, with an average of 50–100 daily visits, making it an ideal site for studying hypertension in this population.

### Study population

This study enrolled PLHIV aged ≥18 years, on ART for >6 months, attending the CTC during the study period. Pregnant women and those on contraceptive pills were excluded due to potential blood pressure confounding effects. Eligibility was confirmed using CTC records.

### Sample size and sampling technique

The sample size was calculated using the Cochran’s formula, based on a previous prevalence of 26% from prevalence from a Tanzanian study [22], with adjustments for 10% non-response rate. The final sample size was 329 participants. Simple random sampling was employed: daily CTC patient lists were generated, each patient assigned a random number, and 329 were selected using a computer-generated random number generator. Of these, 296 participated. Potential recall bias in self-reported data (e.g., smoking, alcohol use, physical activity) was mitigated by structured, pretested questionnaires with clear, simple questions to improve recall accuracy.

### Variables

The dependent variable was hypertension. Independent variables included traditional risk factors (age, gender, smoking, alcohol use, BMI, family history, physical activity), HIV-related factors (duration of HIV diagnosis, CD4 count, WHO stage, ART duration), and knowledge level. There were no missing data for any variables analysed, including sociodemographic characteristics, HIV-related factors, and knowledge of hypertension, as all 296 participants provided complete responses and measurements.

### Data collection procedures

Data were collected from September 10, 2024 to March 29, 2025, through structured questionnaires administered in Swahili during one-on-one interviews in a private room post-clinic visit. Sociodemographic data (age, gender, education, occupation, smoking, alcohol use, physical activity, family history of hypertension) and hypertension knowledge were self-reported. Hypertension knowledge was assessed using a structured 10-item questionnaire adapted from the Hypertension Knowledge-Level Scale (HK-LS) [23], with some of the questions modified to fit the local study context and population under investigation. The tool included questions on: (1) normal blood pressure values; (2–6) behavioural risk factors including cigarette smoking, alcohol consumption, weight gain, excessive salt intake, and lack of physical exercise; and (7–10) complications of hypertension including heart problems, visual disturbances, stroke, and kidney disease. Each correct response was awarded one point and summed to generate a percentage score (0–100%). Categories of general knowledge levels were low (0–30%), moderate (40–70%), and good (80–100%). HIV-related factors (duration of HIV diagnosis, CD4 count, WHO HIV clinical stage, and ART duration) were extracted from CTC records.

#### Blood Pressure Measurement

Blood pressure was measured by two registered nurses using a calibrated sphygmomanometer, following American Heart Association guidelines [24]. Participants rested for 5 minutes, seated with feet flat, arm supported at heart level. Two readings, 1–2 minutes apart, were averaged from two separate occasions. Hypertension was defined as ≥140/90 mmHg or current antihypertensive use.

#### Anthropometric Measurement

Height and weight were measured using a Weight and Height Scale, and BMI was calculated (kg/m^2^) using a mobile application. BMI categories were: underweight (<18.5), normal (18.5–24.9), overweight (25–29.9), obese (≥30).

### Data Analysis

Data were analysed using Statistical Package for the Social Sciences (SPSS) 26. Descriptive statistics (frequencies and crosstabs) were used to summarise participant characteristics and determine the prevalence of hypertension. Knowledge was evaluated through frequencies and composite scores. Bivariate logistic regression analysis was conducted to examine the association between each independent variable and hypertension. Subsequently, variables were included in the multivariate logistic regression model to adjust for potential confounders. Odds ratios (ORs) with 95% confidence intervals (CIs) and p-values <0.05 were considered statistically significant.

## RESULTS

Table 1 presents the sociodemographic and clinical characteristics of the study participants by hypertension status. A total of 296 participants were enrolled in the study. The majority were female (66.6%), and more than one-third were aged 45–54 years (34.8%). Most participants had attained primary education (66.9%) and were business owners (48.6%), while one-third were unemployed (33.1%). Regarding lifestyle characteristics, 7.8% participants reported smoking cigarettes, 23.6% reported alcohol consumption, and the majority were either not very active or moderately active (89.8%). A family history of hypertension was reported by 21.3% participants. More than half of the study population was overweight or obese 58.8%.

**Table 1:** Participant Characteristics with Hypertension status.

| Characteristic | Total n (%) | Hypertensive n (%) | Non-hypertensive n (%) |
| --- | --- | --- | --- |
| **Overall** | 296 (100) | 97 (32.8) | 199 (67.2) |
| **Age (years)** |  |  |  |
| 18–24 | 12 (4.1) | 2 (16.7) | 10 (83.3) |
| 25–34 | 32 (10.8) | 6 (18.8) | 26 (81.3) |
| 35–44 | 85 (28.7) | 21 (24.7) | 64 (75.3) |
| 45–54 | 103 (34.8) | 32 (31.1) | 71 (68.9) |
| ≥55 | 64 (21.6) | 36 (56.3) | 28 (43.8) |
| **Gender** |  |  |  |
| Female | 197 (66.6) | 64 (32.5) | 133 (67.5) |
| Male | 99 (33.4) | 33 (33.3) | 66 (66.7) |
| **Level of education** |  |  |  |
| No formal education | 22 (7.4) | 13 (59.1) | 9 (40.9) |
| Primary education | 198 (66.9) | 60 (30.3) | 138 (69.7) |
| Secondary education | 58 (19.6) | 18 (31.0) | 40 (69.0) |
| High school | 4 (1.4) | 0 (0.0) | 4 (100.0) |
| College degree | 14 (4.7) | 6 (42.9) | 8 (57.1) |
| **Occupation** |  |  |  |
| Business owner | 144 (48.6) | 38 (26.4) | 106 (73.6) |
| Employed | 46 (15.5) | 15 (32.6) | 31 (67.4) |
| Farmer | 8 (2.7) | 3 (37.5) | 5 (62.5) |
| Unemployed | 98 (33.1) | 41 (41.8) | 57 (58.2) |
| **Lifestyle factors** |  |  |  |
| Cigarette smoker | 23 (7.8) | 3 (13.0) | 20 (87.0) |
| Alcohol drinker | 70 (23.6) | 25 (35.7) | 45 (64.3) |
| **Physical activity** |  |  |  |
| Not very active | 133 (44.9) | 52 (39.1) | 81 (60.9) |
| Moderately active | 133 (44.9) | 38 (28.6) | 95 (71.4) |
| Very active | 30 (10.1) | 7 (23.3) | 23 (76.7) |
| **Family history of hypertension** |  |  |  |
| Yes | 63 (21.3) | 27 (42.9) | 36 (57.1) |
| No | 233 (78.7) | 70 (30.0) | 163 (70.0) |
| **BMI (kg/m<sup>2</sup>)** |  |  |  |
| Underweight (<18.5) | 15 (5.1) | 5 (33.3) | 10 (66.7) |
| Normal (18.5–24.9) | 107 (36.1) | 25 (23.4) | 82 (76.6) |
| Overweight (25–29.9) | 102 (34.5) | 34 (33.3) | 68 (66.7) |
| Obese (≥30) | 72 (24.3) | 33 (45.8) | 39 (54.2) |

The overall prevalence of hypertension was 32.8%. Hypertension prevalence increased with age, with the highest proportion observed among participants aged 55 years and above (56.3%). Higher prevalence was also observed among participants with no formal education (59.1%), unemployed individuals (41.8%), those who were physically inactive (39.1%), those with a family history of hypertension (42.9%), and obese participants (45.8%).

Knowledge was moderate (mean 53.85%), with 26.4% scoring low (0–30%), 52% moderate (40–70%), and 21.6% good (80–100%) (Figure 1).

**Figure 1:**
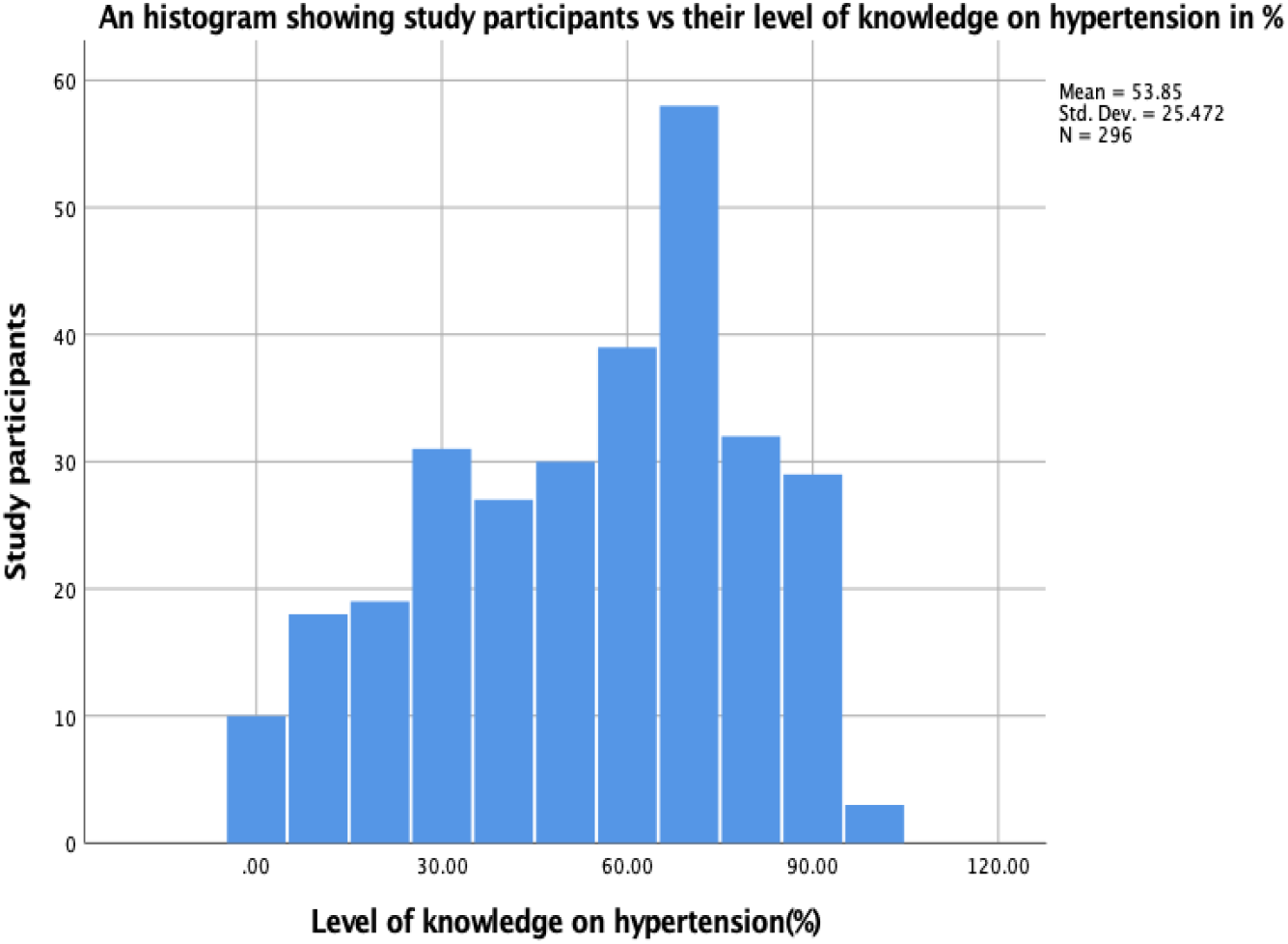
Histogram showing the distribution of hypertension knowledge scores (%)

Only 10.8% knew normal blood pressure, while awareness of risk factors like exercise (90.9%) and weight gain (73.0%) was higher than smoking (22.6%). Complications awareness ranged from 46.3% (kidney problems) to 68.6% (heart problems) (Table 2)

**Table 2:**
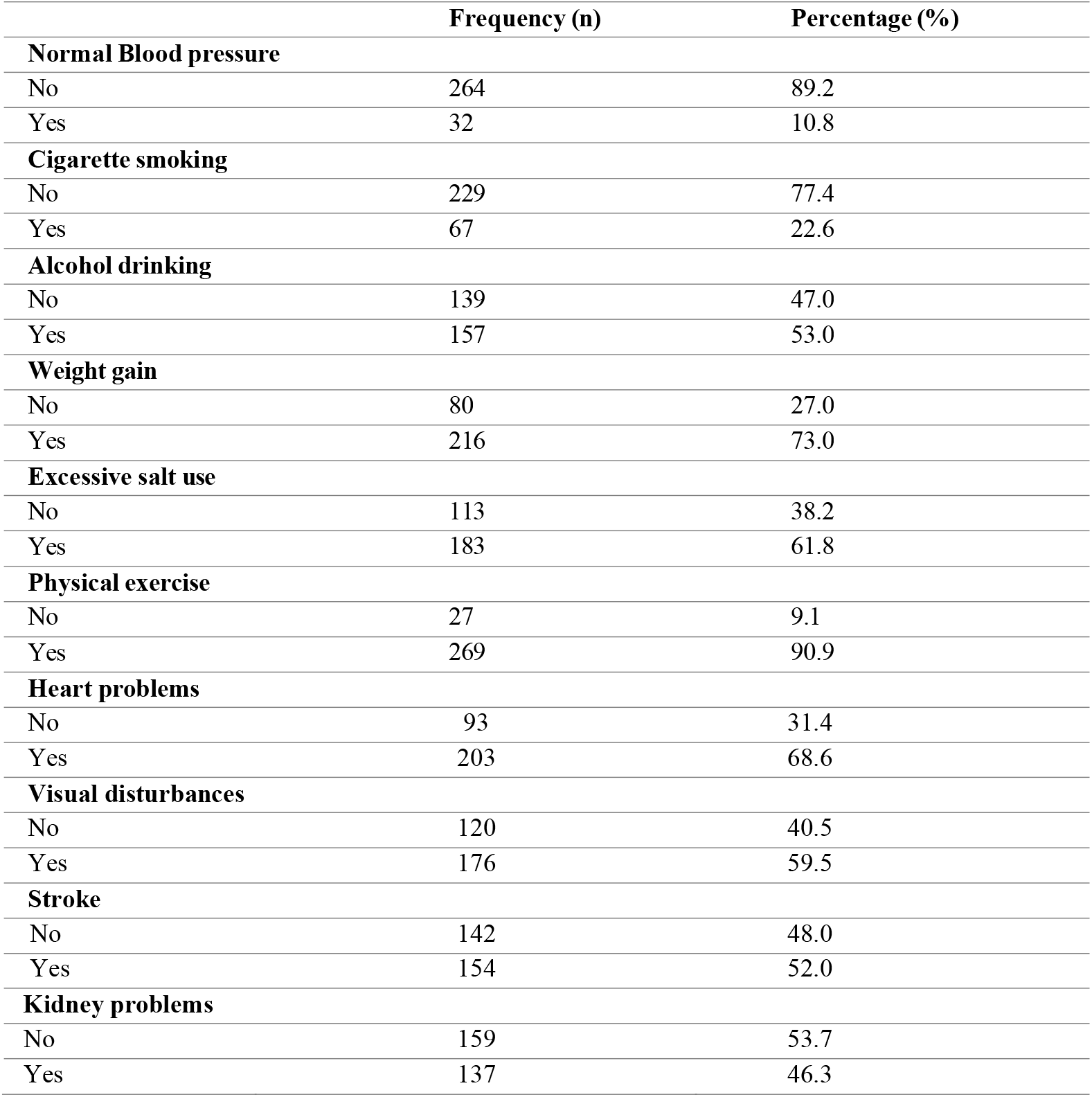
Hypertension knowledge assessment parameters in PLHIV on ART.

|  | Frequency (n) | Percentage (%) |
| --- | --- | --- |
| **Normal Blood pressure** |  |  |
| No | 264 | 89.2 |
| Yes | 32 | 10.8 |
| **Cigarette smoking** |  |  |
| No | 229 | 77.4 |
| Yes | 67 | 22.6 |
| **Alcohol drinking** |  |  |
| No | 139 | 47.0 |
| Yes | 157 | 53.0 |
| **Weight gain** |  |  |
| No | 80 | 27.0 |
| Yes | 216 | 73.0 |
| **Excessive salt use** |  |  |
| No | 113 | 38.2 |
| Yes | 183 | 61.8 |
| **Physical exercise** |  |  |
| No | 27 | 9.1 |
| Yes | 269 | 90.9 |
| **Heart problems** |  |  |
| No | 93 | 31.4 |
| Yes | 203 | 68.6 |
| **Visual disturbances** |  |  |
| No | 120 | 40.5 |
| Yes | 176 | 59.5 |
| **Stroke** |  |  |
| No | 142 | 48.0 |
| Yes | 154 | 52.0 |
| **Kidney problems** |  |  |
| No | 159 | 53.7 |
| Yes | 137 | 46.3 |

Table 3 presents the results of the bivariate and multivariate analyses of factors associated with hypertension among people living with HIV (PLHIV). In the bivariate analysis, age ≥55 years was significantly associated with increased likelihood of hypertension (COR 3.173, 95% CI 2.325–6.395, p<0.001). Cigarette smoking increased the odds of hypertension (COR 3.501, 95% CI 1.014–6.085, p=0.047), while being overweight/obese was associated with lower odds (COR 0.521, 95% CI 0.312–0.870, p=0.013).

**Table 3:**
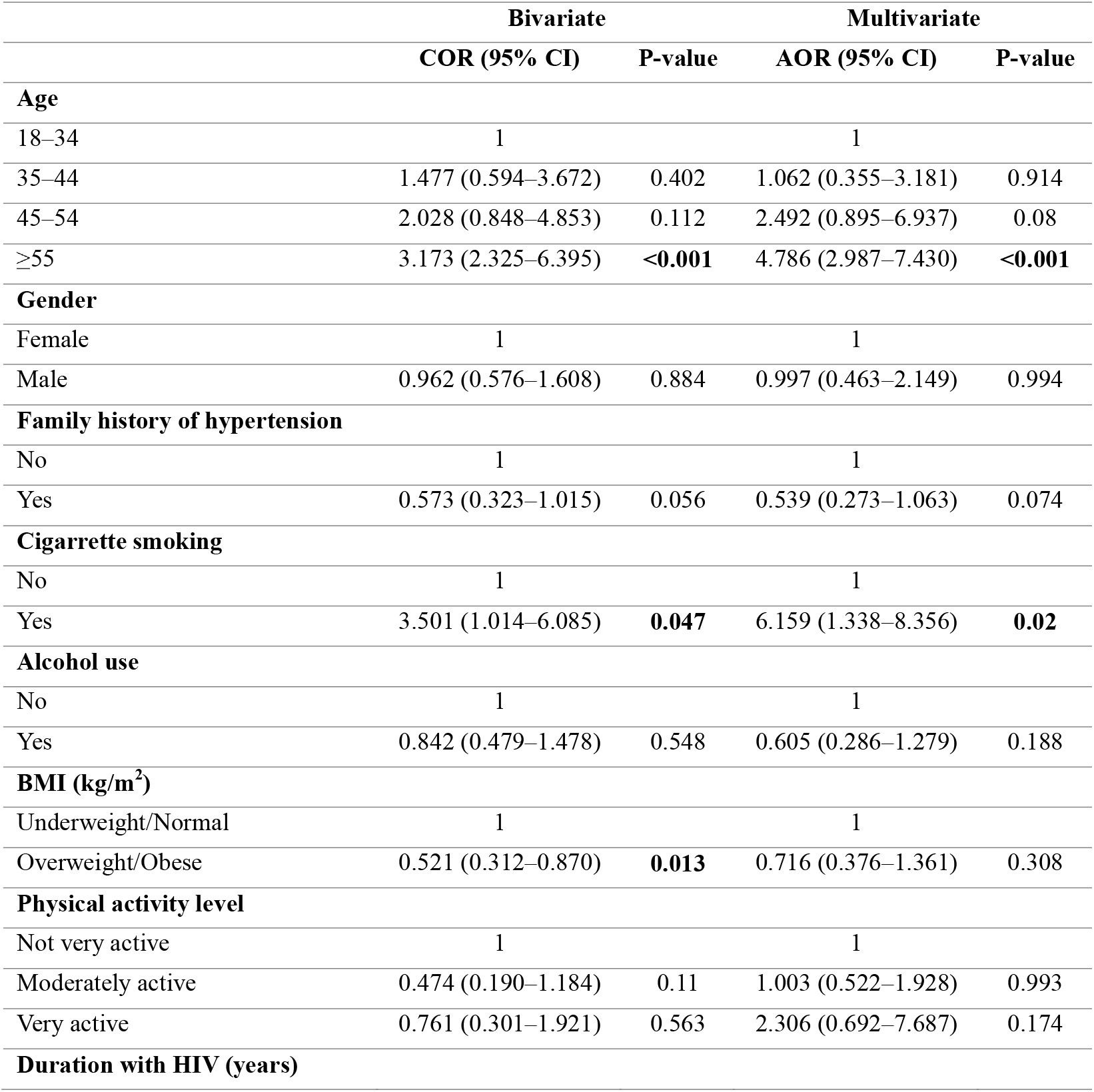
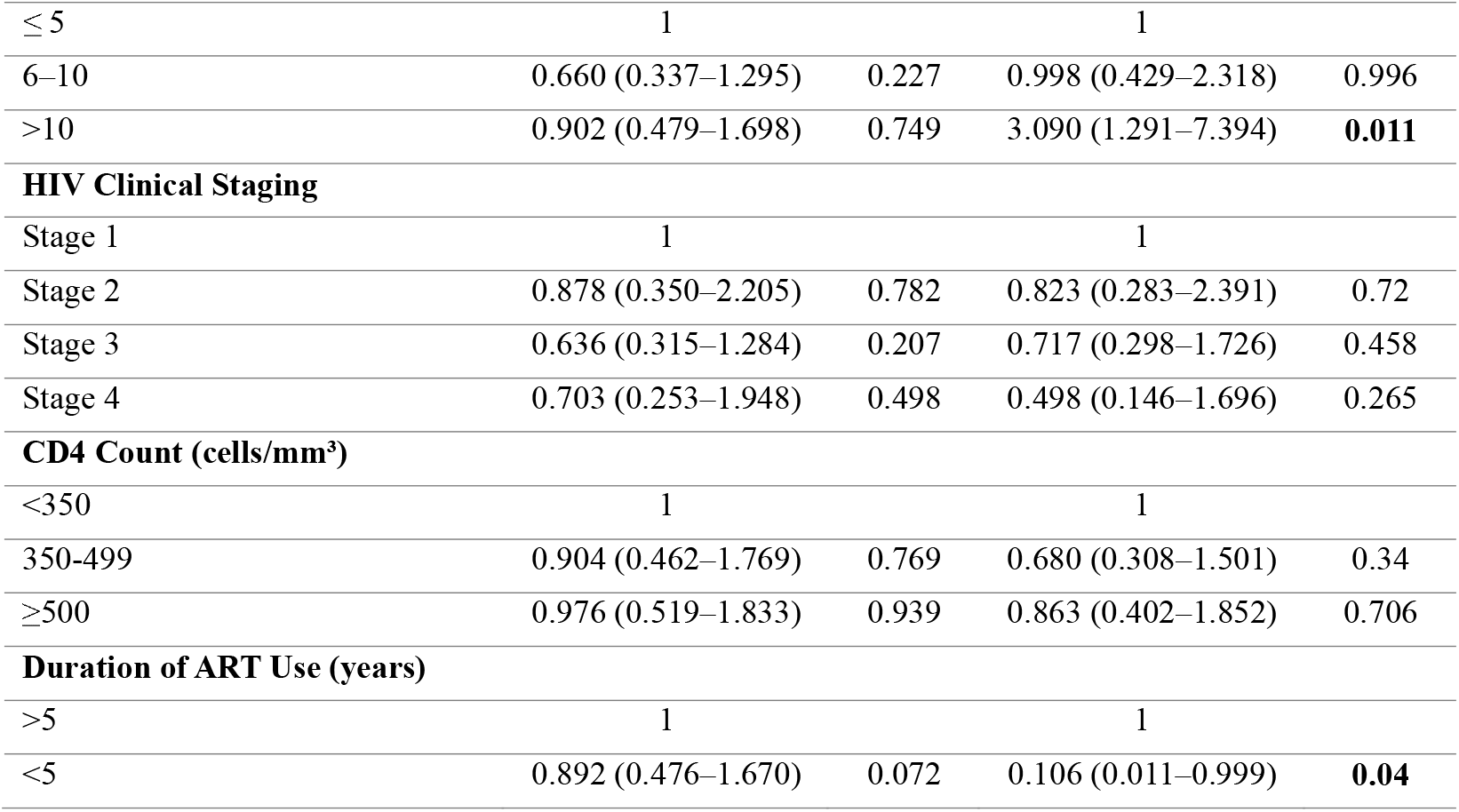
Bivariate and Multivariate analysis of HIV-related factors associated with HTN (N=296)

|  | Bivariate |  | Multivariate |  |
| --- | --- | --- | --- | --- |
|  | COR (95% CI) | P-value | AOR (95% CI) | P-value |
| **Age** |  |  |  |  |
| 18–34 | 1 |  | 1 |  |
| 35–44 | 1.477 (0.594–3.672) | 0.402 | 1.062 (0.355–3.181) | 0.914 |
| 45–54 | 2.028 (0.848–4.853) | 0.112 | 2.492 (0.895–6.937) | 0.08 |
| $\geq 55$ | 3.173 (2.325–6.395) | **<0.001** | 4.786 (2.987–7.430) | **<0.001** |
| **Gender** |  |  |  |  |
| Female | 1 |  | 1 |  |
| Male | 0.962 (0.576–1.608) | 0.884 | 0.997 (0.463–2.149) | 0.994 |
| **Family history of hypertension** |  |  |  |  |
| No | 1 |  | 1 |  |
| Yes | 0.573 (0.323–1.015) | 0.056 | 0.539 (0.273–1.063) | 0.074 |
| **Cigarette smoking** |  |  |  |  |
| No | 1 |  | 1 |  |
| Yes | 3.501 (1.014–6.085) | **0.047** | 6.159 (1.338–8.356) | **0.02** |
| **Alcohol use** |  |  |  |  |
| No | 1 |  | 1 |  |
| Yes | 0.842 (0.479–1.478) | 0.548 | 0.605 (0.286–1.279) | 0.188 |
| **BMI (kg/m<sup>2</sup>)** |  |  |  |  |
| Underweight/Normal | 1 |  | 1 |  |
| Overweight/Obese | 0.521 (0.312–0.870) | **0.013** | 0.716 (0.376–1.361) | 0.308 |
| **Physical activity level** |  |  |  |  |
| Not very active | 1 |  | 1 |  |
| Moderately active | 0.474 (0.190–1.184) | 0.11 | 1.003 (0.522–1.928) | 0.993 |
| Very active | 0.761 (0.301–1.921) | 0.563 | 2.306 (0.692–7.687) | 0.174 |
| **Duration with HIV (years)** |  |  |  |  |
| $\leq 5$ | 1 | | 1 | |
| 6–10 | 0.660 (0.337–1.295) | 0.227 | 0.998 (0.429–2.318) | 0.996 |
| >10 | 0.902 (0.479–1.698) | 0.749 | 3.090 (1.291–7.394) | **0.011** |
| **HIV Clinical Staging** |  |  |  |  |
| Stage 1 | 1 |  | 1 |  |
| Stage 2 | 0.878 (0.350–2.205) | 0.782 | 0.823 (0.283–2.391) | 0.72 |
| Stage 3 | 0.636 (0.315–1.284) | 0.207 | 0.717 (0.298–1.726) | 0.458 |
| Stage 4 | 0.703 (0.253–1.948) | 0.498 | 0.498 (0.146–1.696) | 0.265 |
| **CD4 Count (cells/mm<sup>3</sup>)** |  |  |  |  |
| <350 | 1 |  | 1 |  |
| 350–499 | 0.904 (0.462–1.769) | 0.769 | 0.680 (0.308–1.501) | 0.34 |
| $\geq 500$ | 0.976 (0.519–1.833) | 0.939 | 0.863 (0.402–1.852) | 0.706 |
| **Duration of ART Use (years)** |  |  |  |  |
| >5 | 1 |  | 1 |  |
| <5 | 0.892 (0.476–1.670) | 0.072 | 0.106 (0.011–0.999) | **0.04** |

After adjusting for potential confounders in the multivariate analysis, age ≥55 years remained significantly associated with increased odds of hypertension (aOR 4.786, 95% CI 2.987–7.430, p<0.001). Cigarette smoking was independently associated with increased odds of hypertension (aOR 6.159, 95% CI 1.338–8.356, p=0.020). Duration with HIV >10 years was significantly associated with higher odds of hypertension (aOR 3.090, 95% CI 1.291– 7.394, p=0.011) and ART use for less than 5 years was associated with reduced odds of hypertension (aOR 0.106, 95% CI 0.011–0.999, p=0.040). Other variables, including gender, family history of hypertension, alcohol use, BMI, physical activity level, HIV clinical stage, and CD4 count, were not significantly associated with hypertension (p>0.05).

## DISCUSSION

This study determined the prevalence of hypertension and its associated factors among people living with HIV (PLHIV) receiving antiretroviral therapy (ART). The overall prevalence of hypertension in this study was 32.8%, which is markedly higher than the 11% reported in the general Tanzanian adult population, suggesting a considerable burden of hypertension among PLHIV [16]. This prevalence is higher than the reported global prevalence of hypertension among PLHIV, estimated at 23.6% [10], but falls within the wide range reported across Sub-Saharan Africa (19.6–53%) [11, 12], a region increasingly characterized by the dual burden of infectious and non-communicable diseases [25]. Compared to previous studies conducted in urban Tanzania, which reported hypertension prevalence estimates of approximately 25.2% [20] and 34.6% [16], the findings of the present study indicate a persistently high burden of hypertension in this population. The observed prevalence is comparable to estimates reported in Ghana (36.7%) [21] and South Africa (53%) [12], yet substantially higher than those reported in Burundi (17.4%) [13] and Kenya (16%) [18]. These variations across studies may reflect differences in population age distribution, duration of HIV infection, ART exposure, lifestyle transitions, and the capacity of health systems to detect and manage non-communicable diseases. Collectively, these findings highlight hypertension among PLHIV as a significant and emerging public health concern that warrants systematic integration of non-communicable disease screening and management within HIV care programmes.

Participants’ knowledge of hypertension was assessed, revealing moderate overall knowledge levels (53.85%), with more than one-quarter (26.4%) demonstrating low knowledge. These findings are consistent with previous studies reporting limited hypertension awareness among PLHIV [11, 26, 27]. This study employed a structured and validated approach to assessing hypertension knowledge, improving upon prior studies that relied on non-standardized or vaguely defined measures [22]. By capturing specific domains, including knowledge of normal blood pressure thresholds, behavioural risk factors, and potential complications, this approach provides a clearer understanding of existing knowledge gaps. Although this study did not demonstrate a statistically significant association between knowledge scores and hypertension status, inadequate awareness may still have implications for long-term prevention, health-seeking behaviour, and treatment adherence. Therefore, strengthening patient education within HIV clinics remains important, particularly to support integrated prevention and control of non-communicable diseases within routine HIV care.

Concerning factors associated with hypertension among PLHIV, traditional risk factors such as age and cigarette smoking have demonstrated a significant association. Increasing age was strongly associated with higher odds of hypertension, with participants aged ≥55 years having significantly elevated risk of hypertension. This finding is consistent with extensive epidemiological evidence demonstrating that advancing age remains one of the strongest predictors of hypertension in both the general population and among PLHIV [16, 17]. Age-related vascular changes, including arterial stiffness, endothelial dysfunction, and reduced vascular compliance, are well-established mechanisms contributing to elevated blood pressure [28–31]. Among PLHIV, these age-related processes may be further compounded by chronic inflammation and immune activation, potentially accelerating vascular ageing [32]. As life expectancy among PLHIV continues to improve due to ART scale-up, the ageing HIV population may experience a disproportionately higher burden of hypertension and other cardiovascular comorbidities[33]. These findings underscore the need for early cardiovascular risk assessment within HIV care settings.

Cigarette smoking was also associated with increased odds of hypertension. This finding aligns with previous studies conducted among PLHIV in Sub-Saharan Africa and other settings [9, 34], where smoking has been identified as an important modifiable cardiovascular risk factor. The hypertensive effect of smoking may be mediated through sympathetic nervous system activation, endothelial injury, oxidative stress, and increased arterial stiffness[35, 36]. In PLHIV, the combined effects of smoking and HIV-related inflammation may synergistically elevate cardiovascular risk. However, some studies have reported that there is no association between smoking and hypertension among PLHIV [37, 38], possibly due to underreporting of smoking behaviour, or limited statistical power to identify such associations, which may be attributed to the small sample size and the low proportion of patients. Nonetheless, the present findings reinforce the importance of integrating smoking cessation counselling into routine HIV care as part of comprehensive cardiovascular risk reduction strategies.

HIV-related factors were also significantly associated with hypertension in this study. Longer duration since HIV diagnosis was independently associated with increased odds of hypertension, consistent with findings from previous studies [39, 40]. Even though not assessed in this study, biological mechanisms such as chronic immune activation, persistent inflammation, and endothelial dysfunction have been proposed in the literature to explain the relationship between long-standing HIV infection and increased risk of vascular damage and elevated blood pressure [41, 42]. However, other studies reported no significant association between HIV duration and hypertension [43, 44], possibly due to differences in study design and variability in definitions for duration of HIV infection.

ART duration also demonstrated a significant relationship with hypertension risk. Participants with shorter ART duration (<5 years) had a lower risk of hypertension compared to those on long-term treatment. This finding is consistent with previous studies indicating that prolonged ART exposure may contribute to hypertension through mechanisms such as drug-related metabolic toxicity, lipodystrophy, insulin resistance, and dyslipidaemia [45–48]. As ART programs continue to extend life expectancy among PLHIV, the long-term cardiometabolic consequences of sustained treatment must be proactively addressed through integrated care models. HIV clinical stage was not significantly associated with hypertension in this study, differing from findings reported by other studies [22, 49]. This may be due to ART’s mitigating effects on disease progression.

This study has several limitations. The cross-sectional design limits causal inference between hypertension and associated factors. But also, a detailed assessment of specific ART regimens was not undertaken. Despite these limitations, the study provides important insights into the burden and determinants of hypertension among PLHIV.

## Conclusions

This study demonstrates that hypertension is a major comorbidity among PLHIV, affecting nearly one-third of individuals. Although both traditional and HIV-related factors were associated with hypertension risk, the observed prevalence and key determinants, particularly older age and cigarette smoking, closely mirror patterns documented in the general urban Tanzanian population. These findings suggest that much of the observed risk may reflect ageing, accumulated exposure time, and prolonged survival in care rather than HIV-specific biological mechanisms alone. As life expectancy among PLHIV continues to improve, their cardiovascular risk profile increasingly resembles that of the broader ageing population within Tanzania’s ongoing epidemiological transition. While HIV-related inflammation and ART-associated metabolic effects remain biologically plausible contributors, the present findings primarily support an epidemiological explanation grounded in traditional risk accumulation. The presence of moderate knowledge levels with notable gaps further underscores the need for enhanced patient education within HIV care settings. Integrating routine hypertension screening, targeted health education, and lifestyle counselling into HIV care and treatment clinics is essential to improve early detection, strengthen chronic disease management, and reduce the growing burden of cardiovascular comorbidities among PLHIV in Tanzania.

## Data Availability

The data supporting the findings of this study are available from the corresponding author upon request.

## LIST OF ABBREVIATIONS

AHA: American Heart Association
AOR: Adjusted Odds Ratio
ART: Antiretroviral Therapy
BMI: Body Mass Index
CD4: Cluster of Differentiation 4
CI: Confidence Interval
COR: Crude Odds Ratio
CTC: Care and Treatment Clinic
CVD: Cardiovascular Diseases
HIV: Human Immunodeficiency Virus
HK-LS: Hypertension Knowledge-Level Scale
MRRH: Mwananyamala Regional Referral Hospital
PLHIV: People Living with Human Immunodeficiency Virus
REC: Research Ethics Committee
SPSS: Statistical Package for the Social Sciences
WHO: World Health Organization

## DECLARATIONS

### Ethical approval and consent to participate

Ethical clearance was obtained from the University of Dar es Salaam Research Ethics Committee (REC) with Ref.No. AB3/12(B), with permissions from regional authorities and MRRH. Written informed consent was obtained from all participants, ensuring confidentiality through coded questionnaires.

### Consent for Publication

Not applicable

### Availability of data and materials

The datasets used and/or analysed during the current study are available from the corresponding author on reasonable request.

### Authors’ contributions

Meshack Morice conceptualized the study, collected and analysed data, and wrote the manuscript. Jacqueline Mgumia supervised the study from the beginning to the end and provided critical revisions to the manuscript

## Acknowledgments

The author (s) thank Mwananyamala Regional Referral Hospital (MRRH) staff for their support, and participants for their consent and time to participate in this study.

